# Index microvascular resistance (IMR)-guided management of heart transplantation (IMR-HT study): Study Protocol

**DOI:** 10.1101/2024.11.22.24317768

**Authors:** Ainhoa Pérez-Guerrero, Jean Paul Vilchez-Tschischke, Luis Almenar Bonet, Jose Luis Diez Gil, Teresa Blasco Peiró, Salvatore Brugaletta, Josep Gomez-Lara, José González Costello, Paula Antuña, Vanesa Alonso Fernández, Fernando Sarnago Cebada, María Dolores García-Cosio, Francisco Hidalgo Lesmes, Amador López Granados, Ramón López-Palop, Iris Paula Garrido, Rosa María Cardenal Piris, Diego Rangel Sousa, Georgina Fuertes Ferre

## Abstract

**Background:** Acute allograft rejection (AAR) is an important cause of morbi-mortality in heart transplant (HT) patients, particularly during the first year. Endomyocardial biopsy (EMB) is the “gold standard” to guide post-heart transplantation treatment. However, it is associated with complications that can be potentially serious. Index of microvascular resistance (IMR) is a specific physiological parameter to measure microvascular function. An increased IMR measured early after HT has been associated with acute cellular rejection (ACR), higher all-cause mortality and adverse cardiac events. As far as we know, no study has evaluated IMR impact on post-HT management (number of EMB performed). Our aim will be to assess if post-HT patient management may be modified based on IMR value.

**Study design:** The IMR-HT study (NCT 06656065) is a multicenter, prospective study that will include post-HT consecutive stable patients undergoing coronary physiological assessment in the first three months and one year. Depending on IMR values the physician will be able to reduce the number of biopsies established in each center protocol.

**Conclusions:** Management after heart transplant (number of biopsies) could be modified depending on IMR values.

## INTRODUCTION

Acute allograft rejection (AAR) is an important cause of morbi-mortality after heart transplant (HT), particularly within the first year (1–3). Advances in immunosuppression, donor heart evaluation, surgical techniques, and post-transplantation care have led to a gradual reduction in AAR and improved survival after HT over time. Endomyocardial biopsy (EMB) is the gold standard method to guide post-HT treatment, as it represents the best tool to identify rejection in orthotopic HT (4–5). However, it is usually repeated up to 5 times during the first year - with some variations depending on each center protocol - and it is potentially associated with serious complications.

Several studies have presented the association between AAR, micro-vasculopathy and cardiac allograft epicardial vasculopathy (CAV) (6–9). Index of microcirculatory resistance (IMR) measured early after heart transplantation has been significantly associated with the risk of acute cellular rejection (ACR), and patients with IMR≥15 have higher risk of AAR during 2 years follow-up (10).

The aim of our study is to evaluate if the use of IMR measured at baseline may be useful in guiding care of patients after HT.

## METHODS

### RATIONALE AND DESIGN

The IMR-HT study is a multicenter, prospective study aimed to guide post-HT management based on IMR. It will enroll consecutive eligible patients who undergo HT in each participation center.

#### Patient selection

Patients undergoing HT will be screened for enrollment. To maximize patient inclusion, we apply broad inclusion criteria and strict exclusion criteria, as specified in **Table 1**. Eligible patients will be informed about the study and will have to provide written informed consent prior to being included. The recruitment begins on May 23^rd^, 2023 and the recruitment period of the study is two years. This study adhered to the principles outlined in the Declaration of Helsinki. The approval was granted by the Ethics Committee for Investigation of Aragon (CEICA). The study has been registered at Clinical-Trials.gov (NCT 06656065).

**Table 1.** Eligibility criteria.

| Inclusion criteria |
| --- |
| Patients $\geq 18$ years old who undergo heart transplantation in participant's centers. |
| Informed consent received and signed for study enrollment. |
| Exclusion criteria |
| Patients with hemodynamic instability after HT, including cardiogenic shock or severe coagulopathy. |
| Patients with acute cellular rejection before intracoronary physiological assessment. |
| Patients with bronchial asthma or bronchopathy with a positive bronchodilation test, which contraindicate the use of adenosine. |
| Patients with epicardial coronary lesions with a resting physiological index $\leq 0.89$ or in hyperemia $\leq 0.80$. |
| Patients unlikely to cooperate in the study or with inability or unwillingness to give informed consent. |

The schedule of enrolment, interventions and assessments and the timeline and of the study are detailed in **Fig 1**.

**Figure 1.**
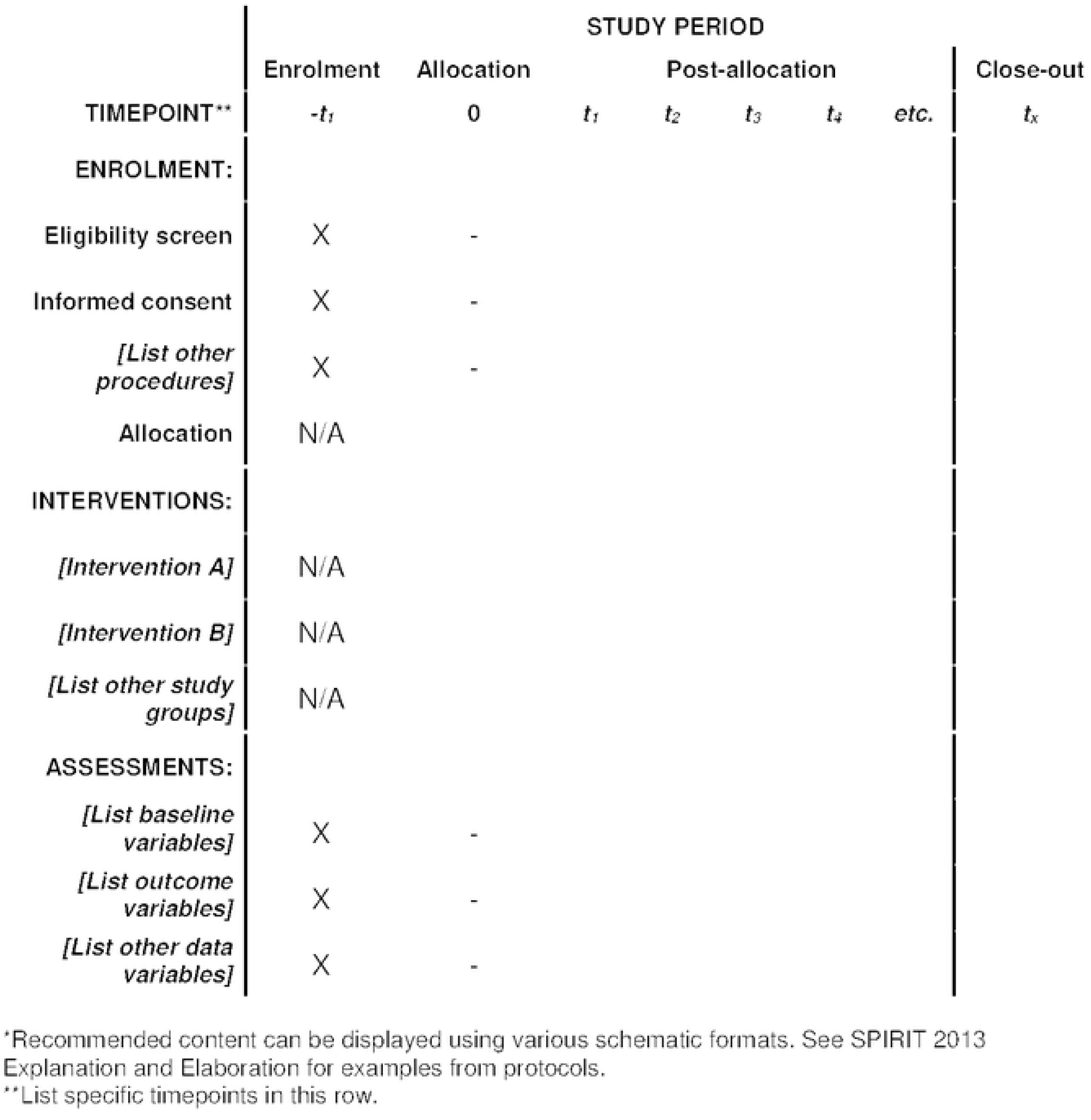
Schedule of enrolment, interventions, and assessments.

Clinical conditions, laboratory findings and clinical events will be assessed at one month and one year. Follow up will be extended for up to five years.

#### Invasive physiological evaluation of the coronary microcirculation

Patients included will undergo a coronary angiography between the first and third month after HT. This coronary angiography is part of the follow-up protocol in most centers also including EMB **(Fig 2).**

**Figure 2.**
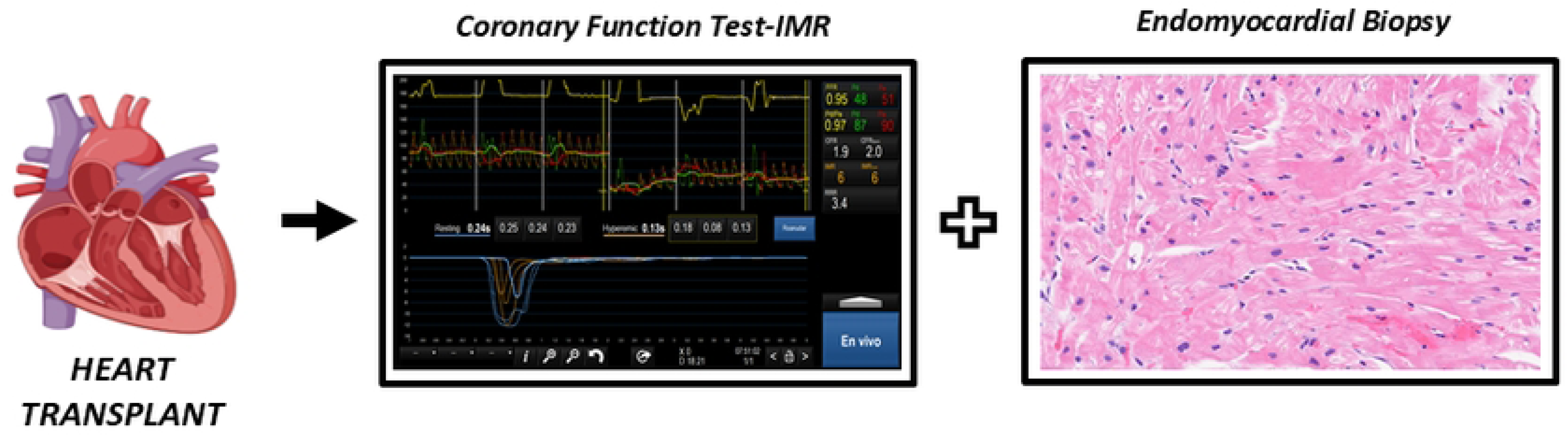
Coronary function test and endomyocardial biopsy between the first and third month after heart transplant.

Assessment of IMR, coronary flow reserve (CFR) and fractional flow reserve (FFR) will be performed using the standard technique (11–13). The left anterior descending coronary artery will be evaluated in all patients. Circumflex or right coronary artery could be additionally evaluated at operator’s discretion. An intracoronary pressure and temperature sensor-tipped guidewire (Pressure Wire TM X guide-wire 0.014’, Abbott, IL, USA) will be used to perform the measurements. The tip pressure sensor will be advanced into the mid-to-distal portion of the evaluated vessel (50 to 60 mm of the ostium of selected coronary artery). Baseline aortic pressure (Pa) and distal intracoronary pressure (Pd) will be obtained to calculate the resting index Pd/Pa. To measure the mean transit time (Tmn) under basal conditions, intracoronary administration of 3 mL of room-temperature saline will be manually injected three times in succession (3 mL/s). Then maximal hyperemia will be induced using adenosine iv (140 to 180 mg/kg/min) and three additional intracoronary room temperature saline boluses of 3 ml will be administered to determine the mean transit time at hyperemia (Tmnh). Deviations >10% in some of the individual Tmn values will force their repetition. Both at rest and in hyperemia, the mean of the three individual determinations will be used for the calculations. Finally, fractional flow reserve (FFR), coronary flow reserve (CFR) and IMR will be calculated using the software Coroventis Coroflow (*Coroventis Abbott, Uppsala, Sweden)*. Physiological indexes are listed in **Table 2**.

- FFR is defined as the ratio of maximal coronary blood flow in a diseased artery to maximal coronary blood flow in the same artery without stenosis. FFR is a surrogate marker of inducible myocardial ischemia caused by epicardial coronary stenosis.
- CFR is the ratio of hyperemic to baseline flow and is a marker of the integrity of both epicardial and microvascular coronary circulation. Therefore, CFR represents the microvascular status when there is no significant epicardial disease.
- IMR is the minimum achievable coronary microcirculatory resistance and a more specific marker of the coronary microcirculation. It is calculated as the ratio of distal coronary pressure to coronary flow at hyperemia and presented in mmHg.s units.

**Table 2.**
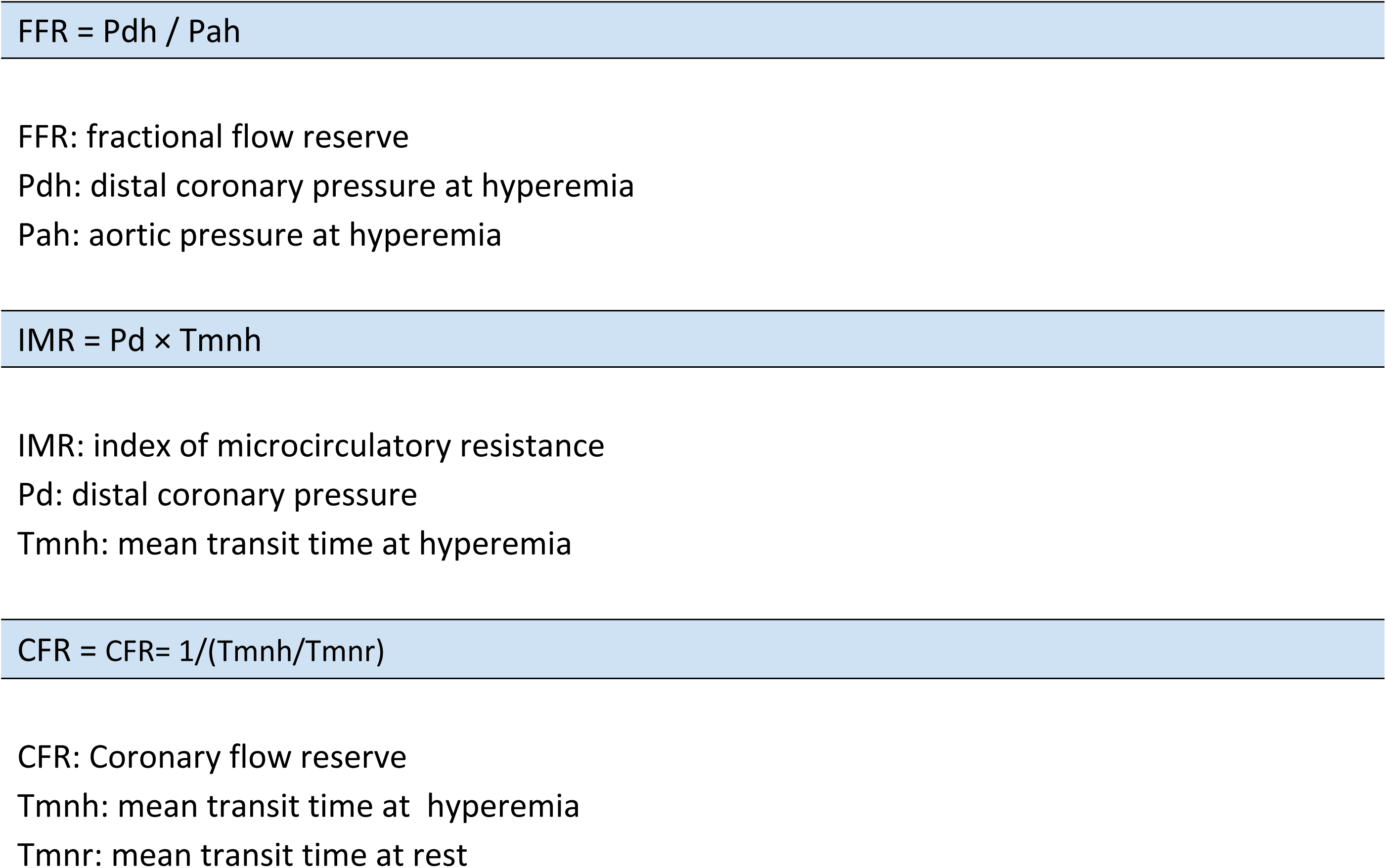
Calculation FFR, IMR and CFR.

The physiological study assessing IMR, CFR and FFR will be repeated again one year after HT.

#### Biopsies and immunosuppressive treatment

Based on the IMR results, the following treatment protocol will be followed:

- If the IMR is less than 15, the number of biopsies could be reduced or kept the same in accordance with the patient’s clinical status and the rest of complementary tests. No immunosuppressive therapy changes would be made. Of note, there may be some physicians that decide not to reduce the number of biopsies despite a low IMR value.
- If the IMR is 15 or greater, immunosuppressive therapy could be intensified or maintained the same. Number of biopsies would be performed as usual per protocol.

The study flowchart is shown in **Fig 3**. Participating centers are listed in Supplementary Table 1.

**Figure 3.**
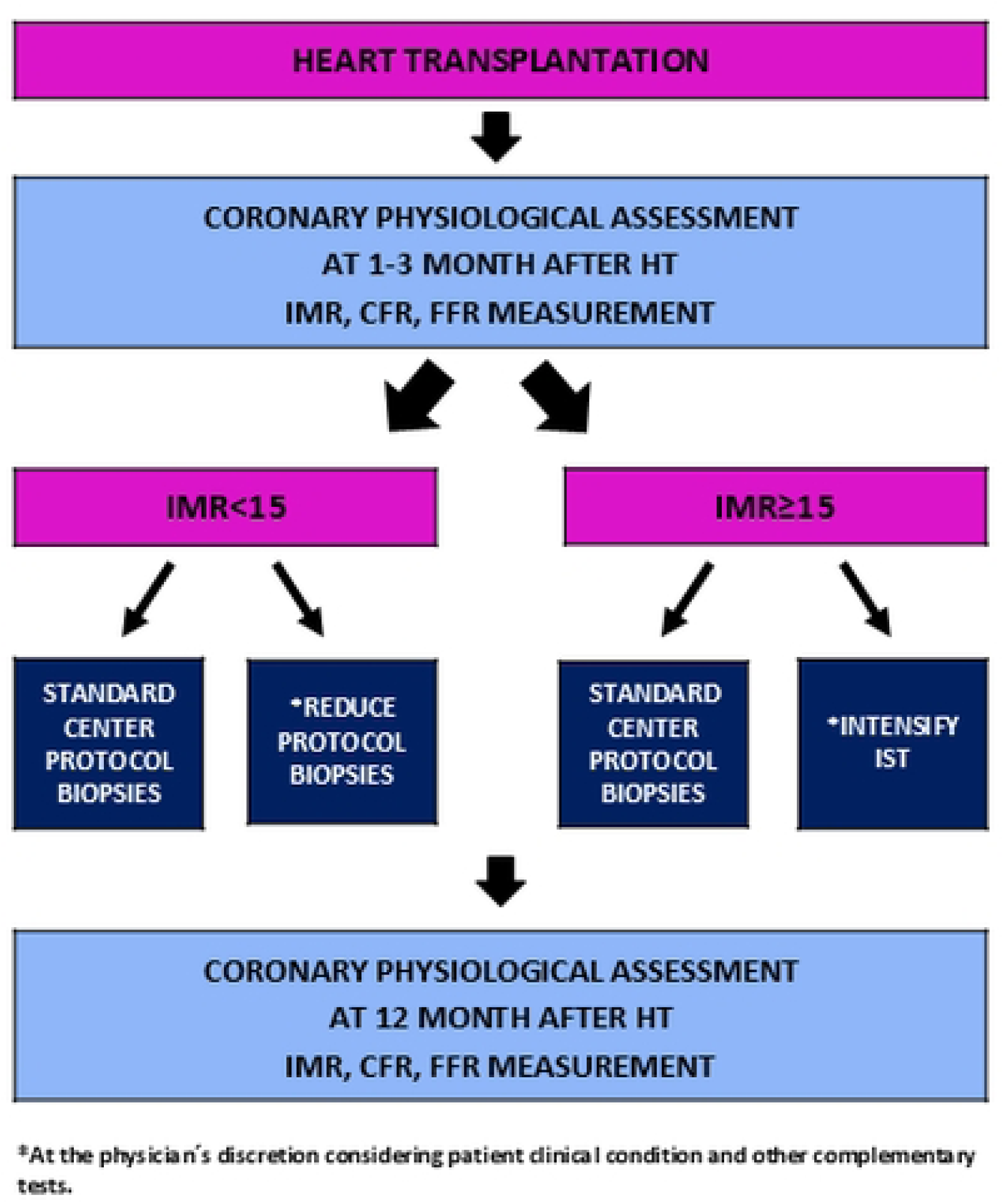
Study Flowchart.

#### Data collection

Sociodemographic, clinical, laboratory and follow-up data of each patient will be included in a database specifically designed for the study. All variables will be included in the online data collection platform Redcap (Research Electronic Data Capture). Each patient will be assigned a number; their identity will not be revealed in any case. All shared information will be anonymized.

#### Study endpoints

The main objective of the study is to evaluate IMR values and number of EMB performed in the first year after HT.

Secondary endpoints are detailed in **Table 3**. End-points will be evaluated at 1 year and annually thereafter for up to 5 years. Definition of events is detailed in **Table 4**.

**Table 3.**
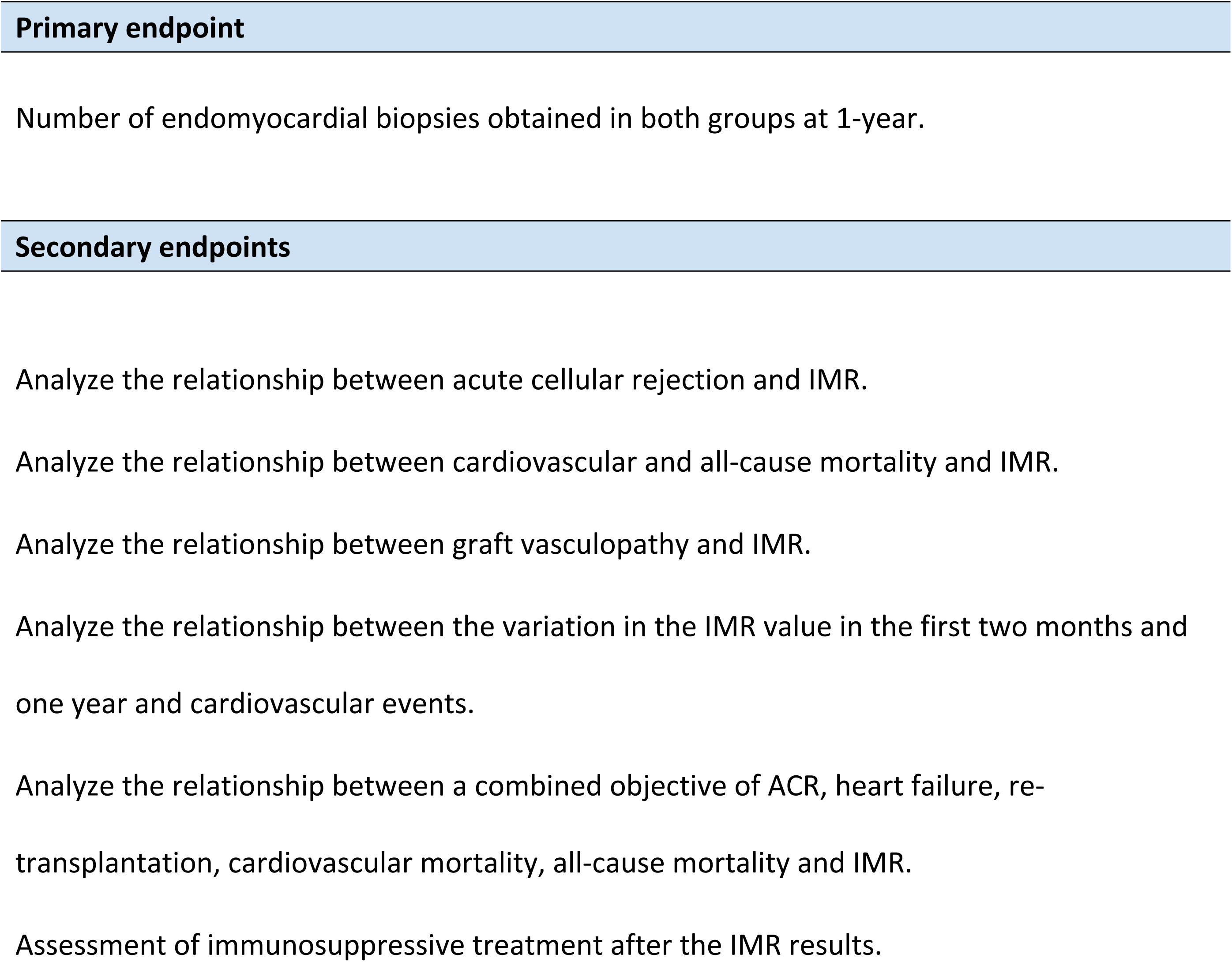
Clinical endpoints.

**Table 4.**
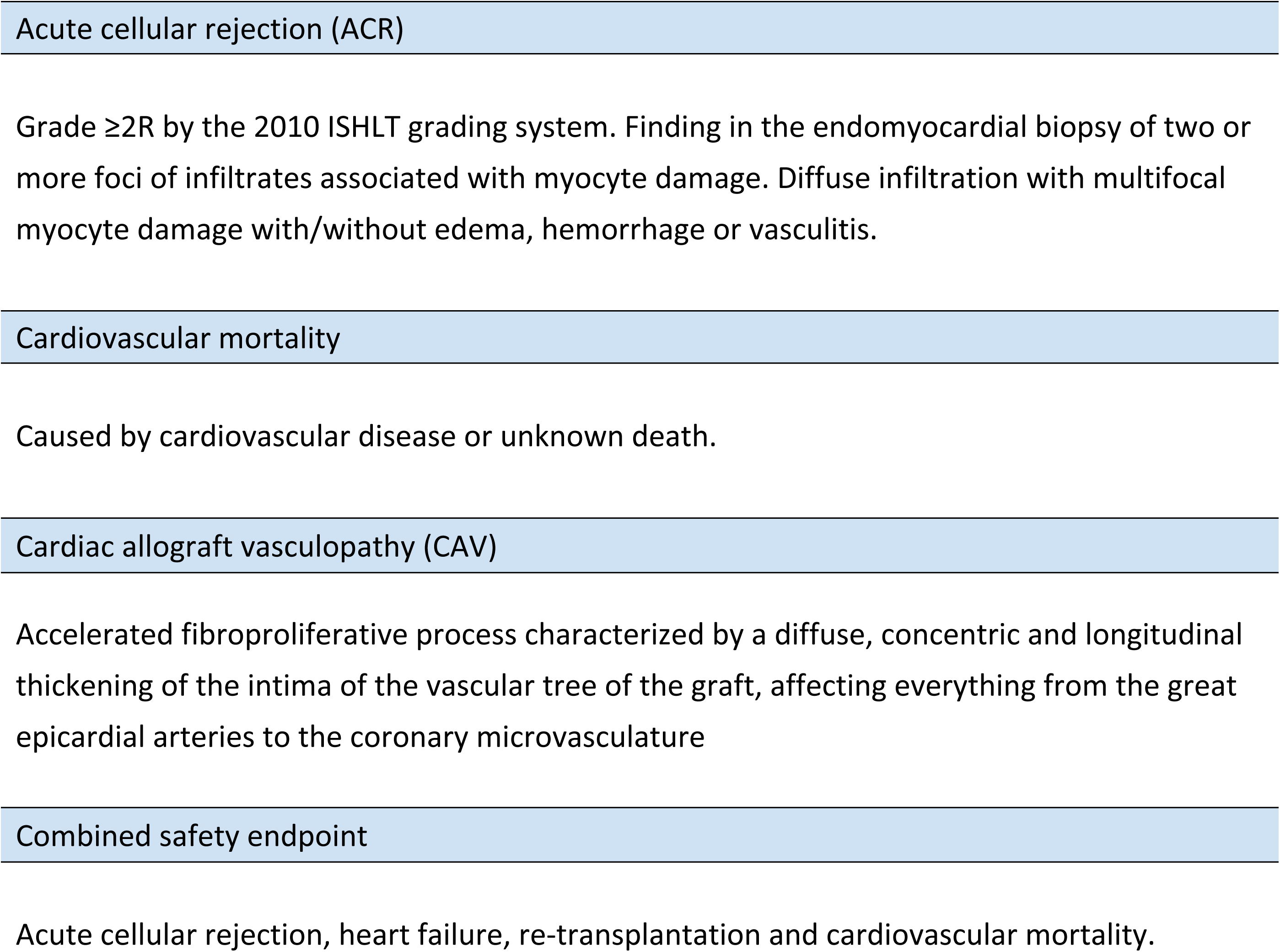
Definition of events.

#### Statistical analysis and sample size considerations

The characteristics of the study population will be summarized through standard descriptive statistics. Continuous variables will be expressed as mean (± standard deviation) or median [interquartile range (IQR)], as appropriate. Discrete variables will be presented as absolute numbers and percentages. For the comparison of means, the Student’s t test for independent measures or the non-parametric Mann-Whitney U test (in the case of dichotomous qualitative variables) and the ANOVA or non-parametric Kruskal test will be used as a hypothesis test. Wallis (in the case of non-dichotomous qualitative variables). For the bivariate analysis of the qualitative variables, the Chi-square test or Fisher’s exact test will be used. Events will be compared between groups by using Kaplan-Meier curves. Cox regression analysis will be performed to adjust for confounding factors and to evaluate independent predictors of clinical events. A two-tailed p-value of 0.05 will be considered statistically significant. All statistical analyses will be performed by SPSS software version 20.

In 2023, around 325 heart transplants were performed in Spain (3). Given a median of 10-15 HT per center and considering that eight centers that routinely perform invasive coronary physiology are participating, data will be analyzed when 100 patients have completed first year follow-up.

## DISCUSSION

Heart transplant is considered the treatment of choice in patients with advanced heart failure refractory to medical treatment or devices (1–2). AAR plays an important role in determining prognosis: up to 20% of HT patients experience at least one episode of ACR in the first year post-transplant (1). The immune response is classified into ACR when it is mediated by T lymphocytes and humoral rejection when the main mechanism involves B lymphocytes and antibody production (4–5, 14). Advances in immunosuppressive therapy (IST), donor heart evaluation, surgical techniques and post-HT care have led to a reduction in ACR, improving survival over time (4). Post-transplant IST includes three basic components: a calcineurin inhibitor (currently preferred Tacrolimus), an antiproliferative agent (mycophenolate mofetil), and steroids. On the other hand, proliferation signal inhibitor (mTOR) drugs (everolimus and sirolimus) are primarily used for CAV (15). CAV is the main cause of mortality after the first year of transplantation. It is characterized by diffuse intimal thickening affecting both coronary epicardial and microcirculation (4,15).

The vast majority of ACR occur asymptomatically, presenting normal ventricular function, and thus being detected through the routine EMB surveillance protocol (4). The graduation of the RAC is detailed in the 2005 review of the ISHLT (14). Due to intra- and inter-observer variabilities in determining different degrees of slight-moderate rejection, an update was published in the 2010 document. In the International Society of Heart and Lung Transplantation (ISHLT) guidelines, EMB was made a IIaC recommendation for the detection of rejection (16). EMB is repeatedly performed during the first year after HT and is associated with complications that, despite infrequent, can be potentially serious, such is the case of cardiac perforation. Moreover, EMB diagnostic yield is wide, with a high variability between observers and a non-negligible rate of false positives and negatives (17).

In order to avoid the inconveniences of EMB, non-invasive techniques have been studied to detect rejection, with some positive results. However, none of these techniques has been able to replace EMB (18,19).

IMR is a quantitative and specific index for coronary microcirculation. An increased IMR in the graft has been associated with higher all-cause mortality and adverse cardiac events regardless of epicardial vasculopathy. Several IMR cut-off (from 15 to >25) have been associated with ACR within the first year after HT. Patients in whom IMR decreases or does not change one year after HT have a higher event-free rate than those patients in whom the IMR increases (8–10, 20–21). However, no study has evaluated IMR impact on post-HT management. In presence of a low IMR value, EMB could be performed less frequently; on the other hand, if the IMR value is high, immunosuppression therapy could be modified by an earlier administration of mTor-inhibiting drugs or prescribing calcium antagonists, which are known to improve microvascular function.

Our aim will be to assess the number of biopsies performed in the first year after baseline IMR results. We believe the results of this trial could be very important for the guidance of future randomized studies.

## CONCLUSIONS

The IMR is a quantitative physiological parameter to evaluate coronary microcirculation. High IMR values have been associated with acute cellular rejection in heart transplant patients. Management after heart transplant (number of biopsies) could be modified depending on IMR values.

## Data Availability

Deidentified research data will be made publicly available when the study is completed and published.

## ABBREVIATIONS

AAR: acute allograft rejection
ACR: acute cellular rejection
CAV: cardiac allograft vasculopathy
CFR: coronary flow reserve
EMB: endomyocardial biopsy
FFR: fractional flow reserve
HT: heart transplant
IMR: index of microcirculatory resistance
IST: immunosuppressive therapy
MVD: microvascular dysfunction
Pa: aortic pressure
Pah: aortic pressure at hyperemia
Pd: distal coronary pressure
Pdh: distal coronary pressure at hyperemia
RFR: resting full-cycle-ratio
Tmnh: mean transit time at hyperemia
Tmnr: mean transit time at rest

## ACKNOWLEDGMENTS

We would like to thank Dr. Mailen Guerrero, from the Anatomical Pathology Department for her contribution to this project.

## SUPPORTING INFORMATION

**S1. SPIRIT checklist**

**S2. Table. Site centers and investigators sample**

